# Stroke recognition in medical emergency calls: A novel sensitivity definition as a basis for developing artificial intelligence decision support

**DOI:** 10.1101/2024.09.24.24314336

**Authors:** Emil Iversen, Hege Ihle-Hansen, Kari Krizak Halle, Alexander Selvikvåg Lundervold, Lars Myrmel, Anders Strand Vestbø, Annette Fromm, Cristian Autenried, Guttorm Brattebø

## Abstract

**Background:** The sensitivity of emergency medical communication centers (EMCC) for stroke detection varies widely. However, few studies offer detailed insights into the entirety of prehospital pathways in patients with stroke. Therefore, this study aimed to lay the foundation for artificial intelligence (AI) decision support tools in EMCCs by exploring their ability to detect strokes in medical emergency calls, describe a novel method for stroke sensitivity calculation in the EMCC, and identify factors associated with stroke recognition during a call.

**Methods:** In total, 1,164 patients with stroke in the catchment area of Bergen EMCC in 2018 and 2019 were included, and a dataset from the EMCC was established manually and linked with data from the Norwegian Stroke Registry (NSR) for analysis. Descriptive statistics, Chi-square test for categorical variables, Mann–Whitney U test for continuous variables, and multivariate logistic regression (LR) were performed on data obtained from patients primarily assessed by EMCC (n=838).

**Results:** Using a novel method, we found a stroke detection sensitivity of 76.8% in our study, compared to the 63.4% when using the traditional sensitivity detection method. LR analysis showed a positive association between stroke suspicion and ischemic strokes (odds ratio [OR]=0.317 [0.209–0.481]; p<0,001, with ischemic stroke as the reference) and wake-up strokes (OR=1.716 [1.110–2.653]; p=0.015). Among the NSR symptoms, only aphasia/dysarthria was positively associated with stroke suspicion (OR=1.600 [1.087–2.353]; p=0.017), while leg paresis (OR=0.609 [0.390–0.953]; p=0.009) and vertigo (OR=0.376 [0.204–0.694]; p=0.002) were negatively associated.

**Conclusions:** This study introduced a novel and more accurate method for calculating EMCC stroke sensitivity, which is relevant for developing decision support tools, such as AI. Moreover, we identified factors of particular interest for future EMCC research that are relevant to developing AI decision-support tools.

**Clinical trials:** https://clinicaltrials.gov/study/NCT04648449

## Introduction

One in four people will have a stroke in their lifetime, and stroke is the third leading cause of combined death and disability worldwide.^1^ Early stroke recognition in medical emergency calls reduces prehospital delay and improves patient outcomes, as stroke treatment is highly time-dependent.^2–5^ However, there is considerable variation in the capability of emergency medical communication centers (EMCC) to recognize stroke in medical emergency calls, with sensitivities ranging from 17.9% to 83.0%.^5–19^ Most studies on EMCC stroke recognition highlight a strong need for initiatives to improve stroke detection.^6,9,13,18^

Recently, artificial intelligence (AI) has been proposed as an EMCC decision support tool.^20,21^ A comprehensive understanding of the current EMCC system is a prerequisite for integrating AI. However, few studies offer detailed insight into the entirety of the prehospital pathways of patients with stroke, and none thoroughly map all patient contact points from symptom onset to hospital arrival.^22–24^

In all studies known to us, the sensitivity of EMCC for stroke recognition is based on stroke-related criterion codes from the protocol used by the medical dispatchers. However, for various reasons, the operators may sometimes choose a non-stroke criterion code. Hence, the sensitivity of EMCC for stroke detection is often based on an incorrect assumption that medical dispatchers always use stroke criterion codes when suspecting a stroke. This may provide a wrong basis for comparing the current EMCC system with an AI-aided one.

The Artificial Intelligence Support in Medical Emergency Calls (AISMEC) project aims to develop a novel AI-based decision support tool for EMCCs. In this study, we aimed to explore the EMCC’s ability to identify strokes in medical emergency calls, develop a novel method for sensitivity calculation, and identify factors associated with stroke recognition during medical emergency calls.

## Methods

This study followed the STROBE reporting guidelines for observational studies (see Supplemental Material). Given the sensitive nature of the data collected for this study, qualified researchers who are trained in human subject confidentiality protocols can request access to the dataset by contacting the Norwegian National Advisory Unit on Emergency Medical Communication at. The corresponding author had full access to all the data in the study and takes responsibility for its integrity and the data analysis.

### Medical emergency calls in Norway

In Norway, medical emergency calls are answered by medical dispatchers (nurses and ambulance personnel) in 16 regional EMCCs. They advise callers and send an ambulance if necessary or refer the callers to an emergency primary care center (EPCC) in the municipality for further assistance if no ambulance is needed. The patients can also call one of the 94 EPCCs directly for non-urgent inquiries. Likewise, the EPCC forwards the inquiry to the EMCC if the patient needs ambulance assistance. All 16 EMCCs use the same dispatch protocol, known as The Norwegian Index for Emergency Medical Assistance (Index), for triage.^25^ The calls are triaged into three degrees of urgency: “acute,” “urgent,” or “regular,” based on the severity of symptoms. Only “acute” triggers an ambulance with the highest priority (“lights and sirens”), while the others are referred to the EPCC, unless there is an obvious need for ambulance transport, but with low priority.^26^

### Study design

#### Participants

This retrospective register-based study included patients discharged with a stroke diagnosis within the catchment area of Bergen EMCC (n=468,000) in Western Norway. The patients were admitted to Haukeland University Hospital or Haraldsplass Deaconess Hospital between January 1, 2018, and December 31, 2019. To ensure the inclusion of all stroke cases, patients were also identified through registration in the mandatory Norwegian Stroke Registry (NSR).^27^ Exclusion criteria were the lack of consent or relevant available data from the Emergency Medical Information System (AMIS). The study was approved by the Regional Committee for Medical Research Ethics Western Norway, REK West (108573) and the Data Protection Officers at Bergen and Haraldsplass Health Trusts (1612-1612). All study participants consented passively to participate in the study and were given the option to withdraw from the study by a postal letter.

### Variables

#### Variables from the NSR

We obtained patient characteristics and stroke-related data from the NSR (see Supplemental Material for details).^28^

The variable “level of consciousness (upon hospital admission)” was dichotomized into “awake” (0) and “others” (1), the latter including all other levels of consciousness (drowsy, adequate reaction to minor stimulation; drowsy, reaction upon major/repeated stimulation; unresponsive or responds with non-targeted movement). We excluded intracerebral hemorrhage when analyzing intravenous thrombolysis (IVT) and endovascular thrombectomy (EVT), as these are irrelevant as treatment options in these cases. The variables “speaking problem” and “dysarthria” were merged into “speaking or dysarthria,” corresponding to the Index. Thus, the patients could have 0–3 Face – Arm – Speech – Time (FAST) symptoms. Stroke severity was categorized on the basis of The American National Institutes of Health Stroke Scale (NIHSS) scores as follows (NIHSS score in parenthesis): very mild (0–2), mild (3-5), moderate (6–14), and severe (15–40).^29^

#### Variables from the Emergency Medical Information System

From the patient record system in Bergen EMCC AMIS (see Supplemental Material for details), we extracted all the prehospital contact points and measures of patients with stroke obtained by the EMS from stroke onset to departure from the scene. All contact points and actions were registered with a time stamp, providing a detailed, chronological dataset of every patient’s prehospital contact points. In case of uncertainties, we used the operator-written free text and audio logs (only patients in the year 2019, due to the lack of audio logs from 2018) for clarification.

#### Data linkage

We established our final dataset by merging patient data from the NSR with AMIS data and our manually registered data.

#### Outcome variable

The manually registered variable “stroke suspected” was defined as “yes” if the ambulance was dispatched as “acute” and the Index criterion code was stroke-related or the text written by the medical dispatcher contained stroke-related information, indicating that the medical dispatcher suspected a stroke. It was also registered as “stroke suspected” if the ambulance was dispatched as “acute,” even without evidence of stroke suspicion (e.g., unconscious patient), as the urgent handling was deemed appropriate given the assignment of an acute degree of urgency.

### Statistical analyses

Results are presented as means or medians, with interquartile ranges and standard deviation for continuous variables and frequencies. Categorical variables are described as absolute numbers and proportions in percentages.

Patient characteristics were analyzed for all participants. To focus solely on the EMCC’s ability to suspect stroke, we excluded patients whose cases had been assessed and triaged by a medical doctor before EMCC contact from further analysis. We compared the “stroke suspected” and “stroke not suspected” subgroups using the Mann–Whitney U test for the continuous variables including age, time of day of contact, onset to first contact, and NIHSS. Categorical variables were compared using the chi-square test.

We explored factors associated with the medical dispatchers suspecting stroke using multivariate logistic regression. We focused on patient-related factors (age, sex, and stroke type [ischemic or hemorrhagic]), symptoms, and operational factors (time of contact [day vs. night], time from onset to first contact, and wake-up stroke). For the stroke-related factors, we excluded “level of consciousness on admittance to the hospital,” as this variable may interfere with the other variables’ effect in the logistic regression analyses.

Results are presented with their respective odd ratios (ORs) and 95% confidence intervals (95% CIs). A p-value of <0.05 was considered statistically significant, and p=0.000 was reported as p<0.001. Data were analyzed using IBM SPSS Statistics for Windows, version 29.0.2.0 (IBM Corporation, Armonk, NY, USA), R Statistical Computing Software, version 2022.07.2+576 (R Foundation for Statistical Computing, Vienna, Austria), and Microsoft® Excel® for Microsoft 365 MSO, version 2308 build 16.0.16731.20542 (Microsoft Corporation, Redmond, WA, USA).

## Results

Of the 1,303 patients enrolled from the NSR, 838 were primarily assessed by a medical dispatcher and included in the analyses. Figure 1 shows the flow chart of the study group selection.

**Figure 1:**
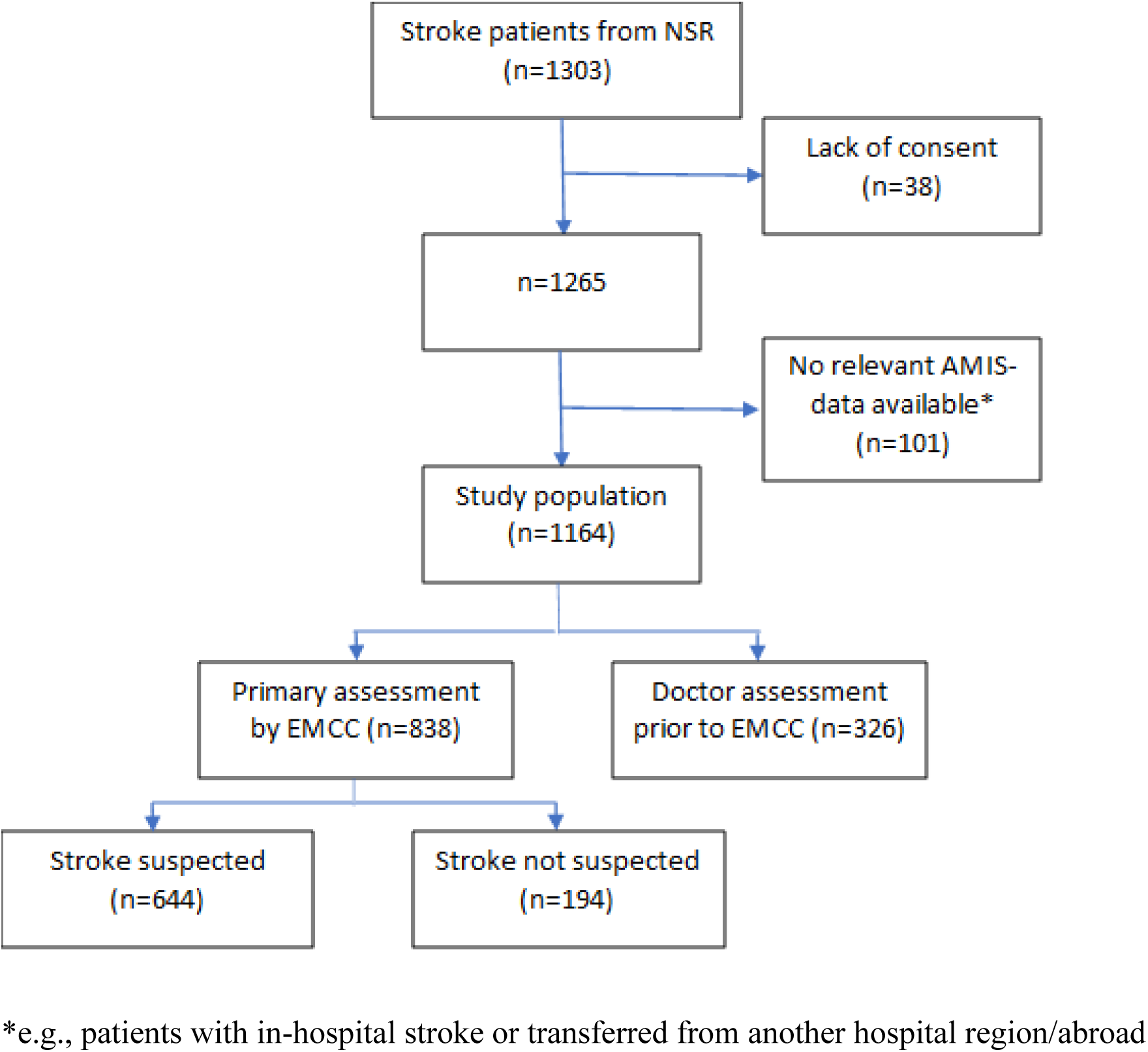
Flowchart of the study population. NSR, Norwegian Stroke Registry; AMIS, Emergency Medical Information System; EMCC, Emergency Medical Communication Centers

### Patient characteristics

Patient characteristics are presented in Table 1. There was no significant difference in age or sex between the “stroke suspected” (n=644) and “stroke not suspected” (n=194) subgroups. However, there was a significantly higher proportion of wake-up strokes, awake patients, and IVT treatment among “stroke suspected” patients. Further, we found a significantly higher proportion of ischemic strokes in the “stroke suspected” subgroup and hemorrhagic strokes in the “stroke not suspected” subgroup. The stroke severity at admission was significantly different, with more cases of “very mild” and “major” strokes in the “stroke not suspected” subgroup and more “mild” and “moderate” stroke cases in the “stroke suspected” subgroup.

**Table 1:**
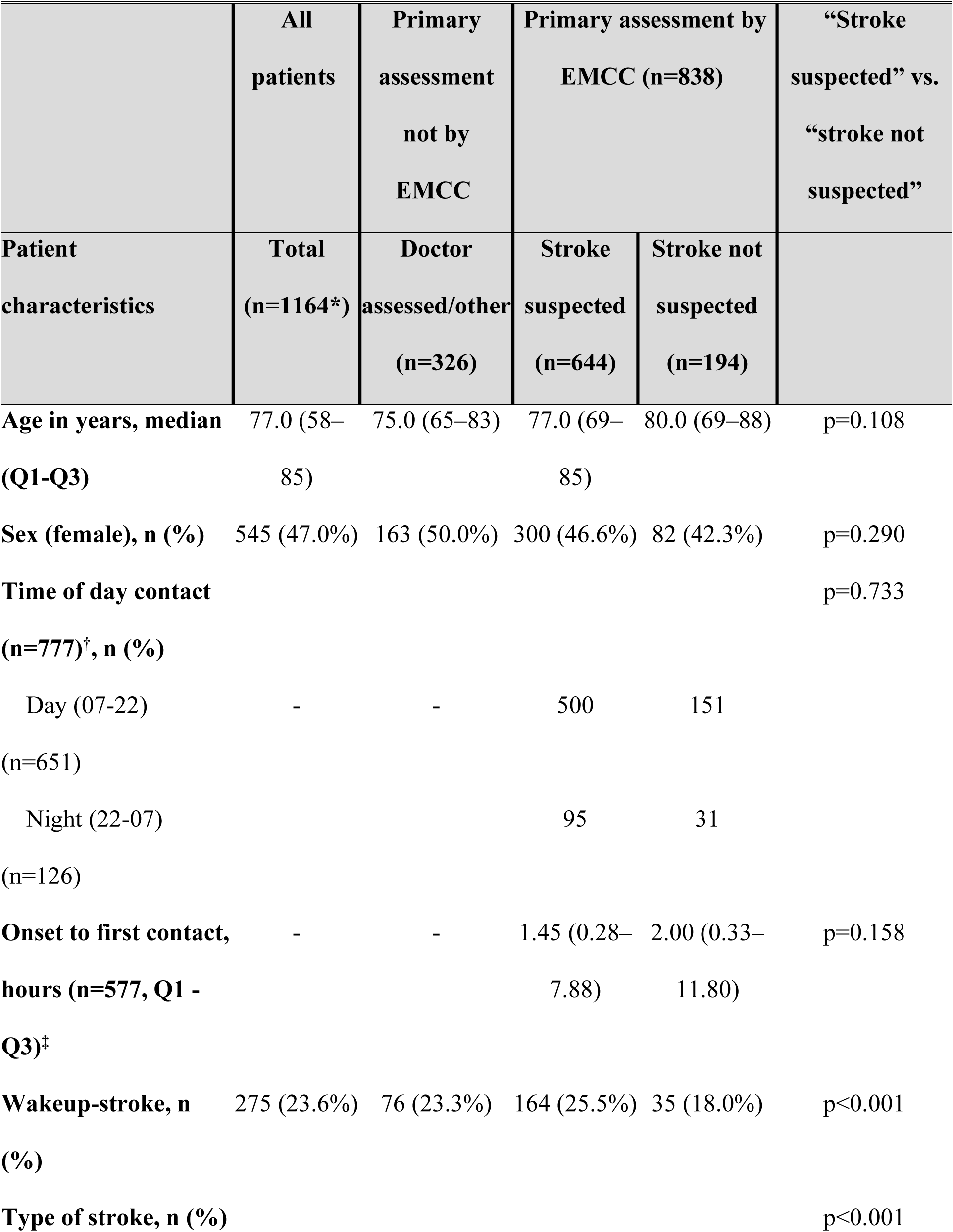
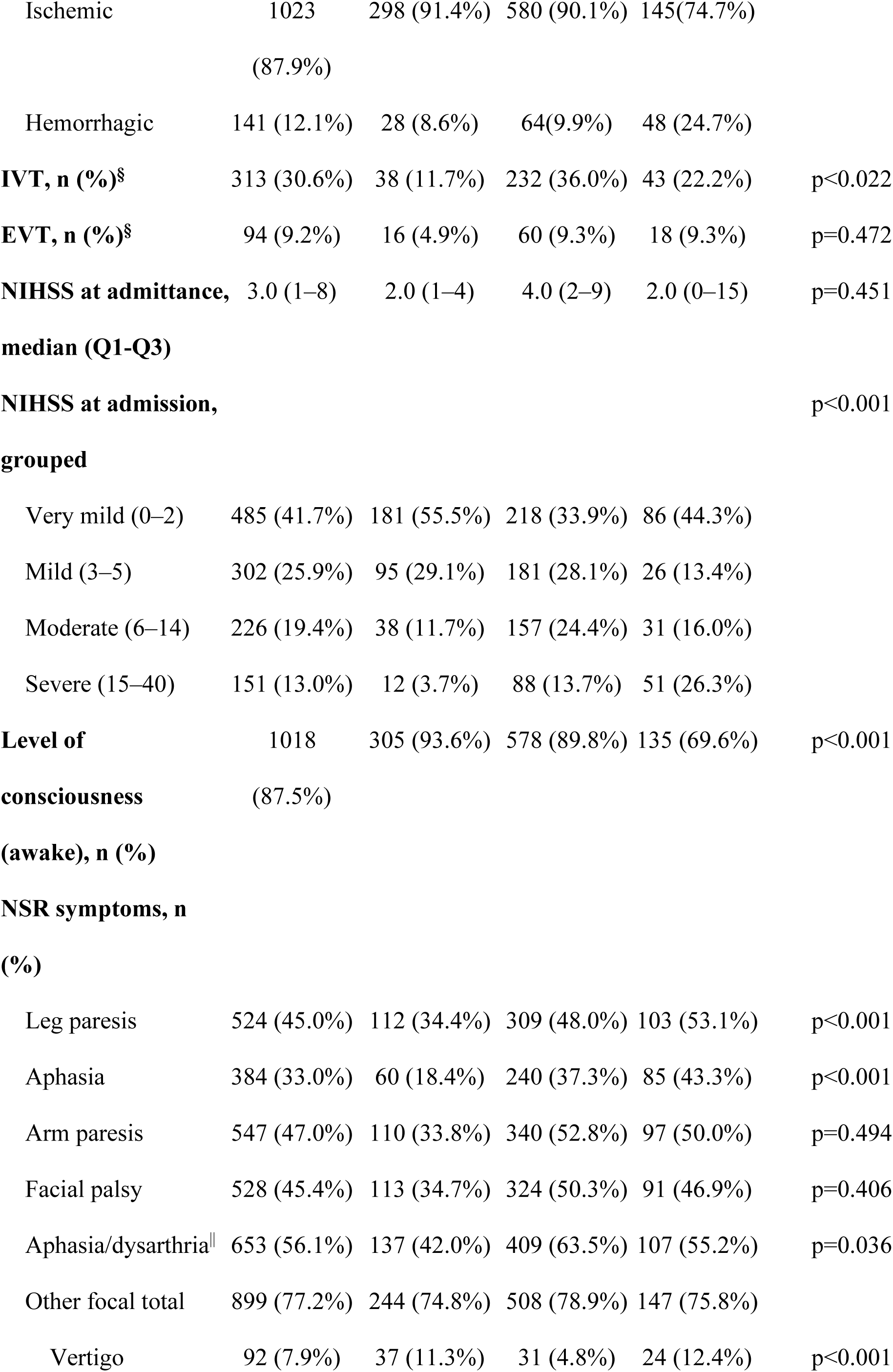
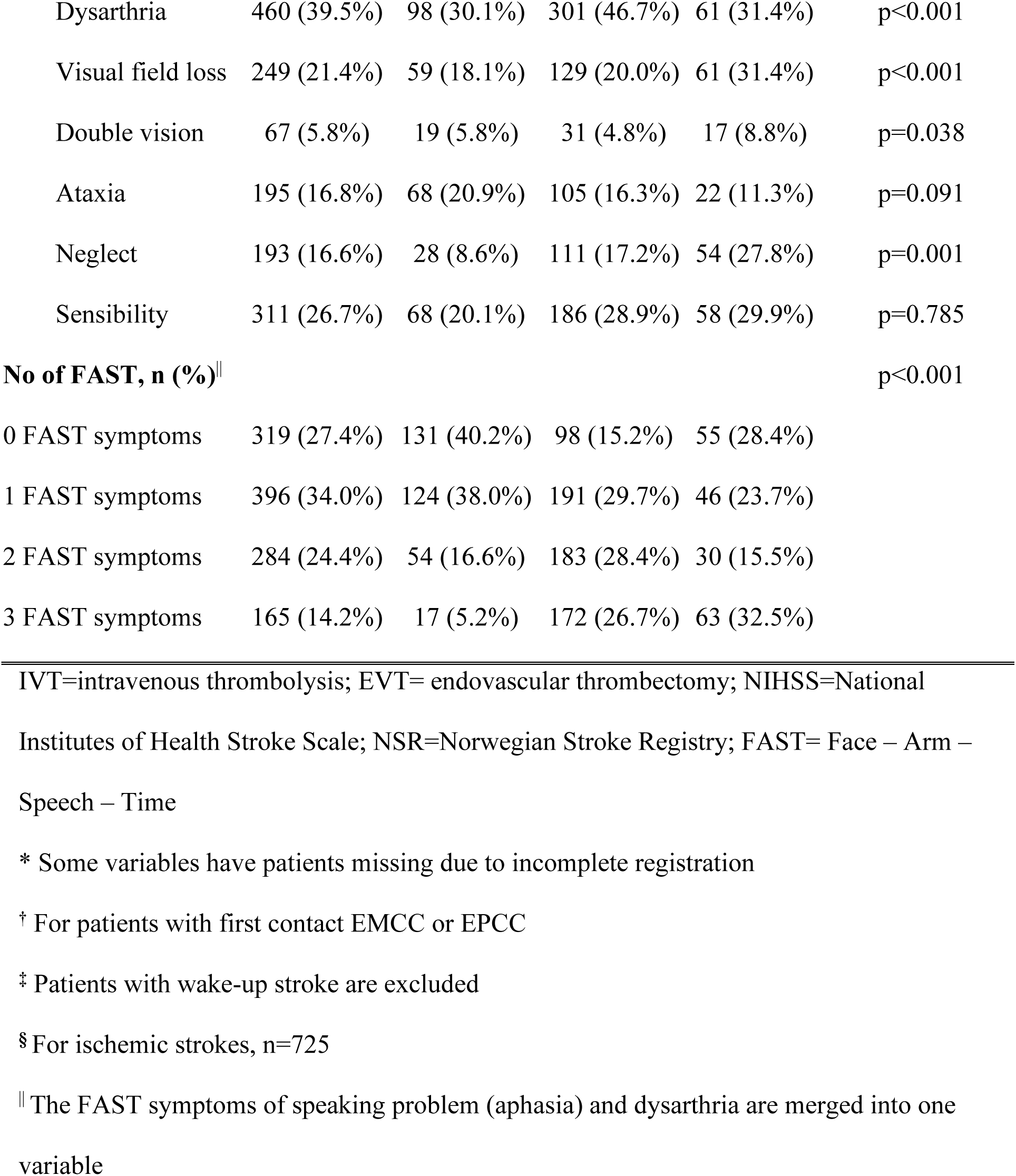
Patient characteristics and comparison of the “stroke suspected” and “stroke not suspected” subgroups.

Leg paresis, visual field loss, double vision, neglect, and vertigo were significantly more frequent in the “stroke not suspected” subgroup than in the “stroke suspected” subgroup. Symptoms, including problems with speaking or dysarthria (aphasia/dysarthria combined), were more prevalent in the “stroke suspected” subgroup than in the other group.

### “Stroke suspected”

In the “stroke suspected” subgroup (n=644), 531 patients (82.5%) were assigned a stroke criterion. In 95 cases (14.8%), the evidence of suspected stroke was in free written text. Finally, 18 (2.8%) of the “stroke suspected” cases were included due to the use of the “acute” criterion without other stroke-related information.

### EMCC stroke sensitivity

Table 2 shows that EMCC stroke sensitivity calculated as normal in traditional EMCC research was 63.4% and 76.8% using dispatch protocol criterion codes and the “stroke suspected” variable, respectively.

**Table 2:** Traditional vs. novel stroke sensitivity by groups for 838 patients primarily assessed by medical dispatchers.

|  | **Yes** | **No** | **Total** | **Sensitivity** |
| --- | --- | --- | --- | --- |
| Assigned Index stroke criteria<br>(traditional sensitivity) | 531 | 307 | 838 | 63.4% |
| “Stroke suspected” by EMCC<br>(novel sensitivity) | 644 | 194 | 838 | 76.8% |

Table 3 shows the use of Index criterion codes for the study participants. In total, 359 (30.8%) patients were assigned 110 different non-stroke criterion codes from 19 (of the total 39) chapters in the Index (see Supplementary Material). There was a significant difference between the groups (p<0.001).

**Table 3:**
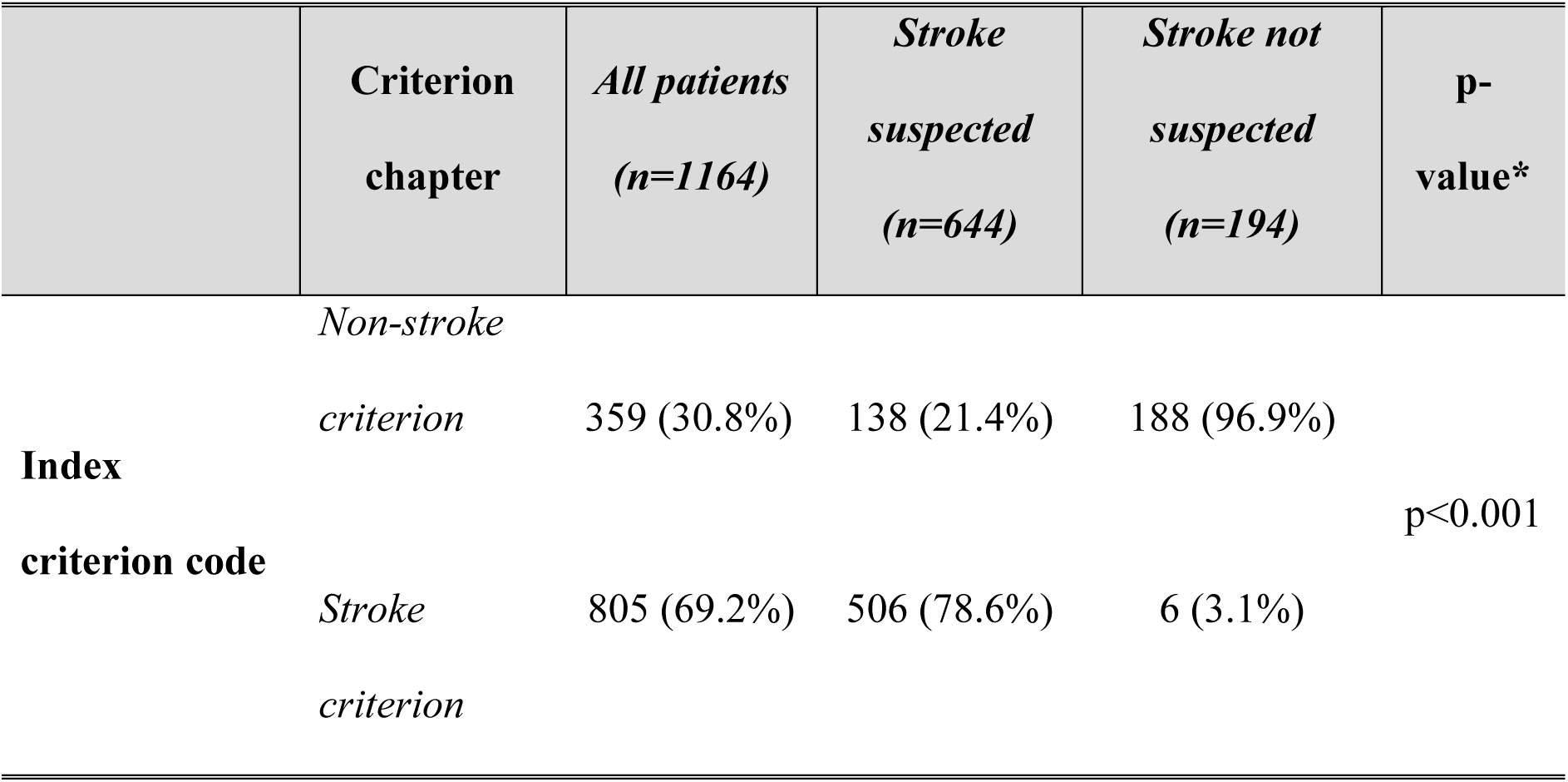
Distribution of stroke vs. non-stroke Index criterion codes for all patients and subgroups.

### Factors associated with stroke suspicion

Table 4 presents the variables associated with the suspicion of stroke by medical dispatchers. We found that neither age nor sex influenced stroke suspicion. However, ischemic stroke (adjusted for age and sex) was significantly associated with stroke suspicion (OR for ischemic vs. bleeding 0.32; 95% CI = 0.21–0.48), indicating a higher likelihood of stroke suspicion for ischemic than hemorrhagic strokes.

**Table 4:** Association between medical dispatchers’ stroke suspicion and patient-related factors, symptoms, and operational factors.

|  | **Coefficient** | **Std.**<br>**error** | **p-**<br>**value** | **Odds**<br>**Ratio** | **95% CI** |
| --- | --- | --- | --- | --- | --- |
| **Patient-related factors** |  |  |  |  |  |
| Age* | -0.01 | 0.01 | 0.141 | 0.99 | 0.98–1.00 |
| Sex* | 0.28 | 0.17 | 0.105 | 1.3 | 0.94–1.86 |
| Stroke type<sup>†</sup> | -1.15 | 0.21 | <0.001 | 0.32 | 0.21–0.48 |
| **Symptoms** |  |  |  |  |  |
| Age* | -0.01 | 0.01 | 0.153 | 0.99 | 0.98–1.00 |
| Sex * | 0.25 | 0.18 | 0.155 | 1.29 | 0.91–1.82 |
| Vertigo | -0.978 | 0.31 | 0.002 | 0.38 | 0.20–0.69 |
| Arm paresis | 0.42 | 0.23 | 0.070 | 1.52 | 0.97–2.39 |
| Ataxia | 0.42 | 0.26 | 0.108 | 1.53 | 0.91–2.56 |
| Double vision | -0.32 | 0.33 | 0.328 | 0.73 | 0.38–1.38 |
| Facial palsy | 0.09 | 0.20 | 0.662 | 1.09 | 0.74–1.62 |
| Field of vision | -0.45 | 0.22 | 0.039 | 0.64 | 0.42–0.98 |
| Leg paresis | -0.50 | 0.23 | 0.030 | 0.61 | 0.39–0.95 |
| Neglect | -0.63 | 0.24 | 0.009 | 0.54 | 0.34–0.85 |
| Aphasia/dysarthria | 0.47 | 0.20 | 0.017 | 1.60 | 1.09–2.35 |
| Sensibility | 0.17 | 0.20 | 0.411 | 1.18 | 0.80–1.75 |
| **Operational factor** |  |  |  |  |  |
| Age* | -0.01 | 0.01 | 0.232 | 0.99 | 0.97–1.01 |
| Sex* | 0.23 | 0.18 | 0.196 | 1.26 | 0.89–1.78 |
| Time of day contact | -0.18 | 0.23 | 0.439 | 0.84 | 0.53–1.31 |
| Time to first contact | 0.00 | 0.00 | 0.916 | 1.00 | 1.00–1.00 |
| Wake-up stroke | 0.54 | 0.22 | 0.015 | 1.72 | 1.11–2.65 |

\* Included as adjustment variables
† Reference category: ischemic

Of the symptom variables (adjusted for age and sex), only aphasia/dysarthria was positively associated with stroke suspicion (OR 1.60; 95% CI = 1.09–2.35). We found negative associations for leg paresis (OR 0.61; 95% CI = 0.39–0.95), neglect (OR 0.54; 95% CI = 0.34–0.85), field of vision disturbance (OR 0.64; 95% CI = 0.41–0.98), and vertigo (OR 0.38; 95% CI = 0.20–0.69).

For the operational variables (adjusted for age and sex), only wake-up stroke showed a significant positive association with stroke suspicion (OR 1.72; 95% CI = 1.11–2.65).

## Discussion

In this study, we aimed to explore the EMCC’s ability to identify strokes in medical emergency calls, develop a novel method for sensitivity calculation, and identify factors associated with stroke recognition during medical emergency calls. In line with the objectives, we propose a novel and more accurate way of calculating EMCC stroke sensitivity. We also identified factors associated with stroke suspicion in patients who were not evaluated by a doctor. Both objectives lay the foundation for comparing the current EMCC system with innovative decision support tools, such as AI.

To our knowledge, this new method for calculating EMCC stroke sensitivity, based on a manual review of prehospital stroke data, has not been previously described. Using the manually created variable “stroke suspected” is the most accurate way of calculating EMCC stroke sensitivity, as approximately 15% of the patients we included in our “stroke suspected” group were not assigned a stroke criterion from the dispatch protocol. When a patient suffers a stroke (or other conditions), the circumstances of the call may reasonably lead the medical dispatcher to choose a non-stroke-related criterion (for example, a patient suffering a stroke and falling or getting involved in a car accident). In such cases, using accident-related instead of stroke-related dispatch protocol criteria is justified. Hence, using the traditional way of calculating sensitivity may be the wrong basis for evaluating the accuracy of EMCC stroke detection. The traditional sensitivity also provides an unprecise basis for comparison when developing and testing novel AI-aided decision support tools. Such decision support aims to assist the medical dispatchers to not overlook a condition; however, the final decision on further handling remains with the medical dispatcher and not the AI.

Thus, accurate sensitivity, based on not only the dispatch protocol criteria code but also the medical dispatchers’ evaluations, is necessary. As highlighted by Yu et al., this process is complex, and there is a close link between the development and use of the AI model and human values, requiring close attention and guidance.^30^ Using a correct method for evaluating the accuracy of stroke detection is one of the basics of this fine-tuned process. The technique can be applied in EMCC systems in any location if a relevant dispatch protocol and patient record system are used in the EMCC.

Notable findings emerged regarding factors associated with stroke suspicion. Specifically, no association was found between age or sex and stroke suspicion. This is in contrast with results of a recent Australian study, which highlighted female sex as a risk factor for wrong diagnosis in the emergency setting.^31^ Cultural, linguistic, or society-related factors can contribute to this discrepancy, as it is hard to imagine different symptom presentations between countries or regions. Regarding stroke type, we found a higher probability of stroke detection for ischemic than for hemorrhagic strokes. This is consistent with the findings of an Australian study, which reported a lower likelihood of stroke suspicion in patients with intracerebral hemorrhage.^10^ However, the practical application of this finding is limited, as accurately determining stroke type during the initial medical emergency call seems to be a distant prospect. Among the stroke symptoms from the NSR, only aphasia/dysarthria was positively associated with stroke suspicion. This might be explained by the fact that it is easier for the caller to describe speech-related problems. Sometimes, the patient places the call, and in such cases, the aphasia/dysarthria is evident to the medical dispatcher. In a Finnish study, speech disturbance was the most frequently overlooked FAST symptom, which was identified as a target for improvement.^32^ Overall, aphasia/dysarthria is a notable symptom in the EMCC setting and warrants further investigation.

Vertigo, leg paresis, neglect, and field of vision disturbance were negatively associated with stroke suspicion. Regarding leg paresis and vertigo, a stroke affecting a patient’s ability to stand or walk might lead to events where the medical dispatcher assigns a non-stroke criterion. This influences the sensitivity of EMCC stroke detection, possibly resulting in a false low sensitivity, as shown in this study.

Additionally, medical dispatchers might fail to recognize a patient suffering a stroke, which could result in not dispatching an ambulance with the highest degree of urgency. This result is consistent with that of previous studies, and falls have been described as the presenting symptom in approximately 38% of inquiries regarding stroke in medical emergency calls.^32,33^ Hence, there is a need for decision support systems that can identify stroke as a potential cause of falls. Neglect, described as a lack of attention, and field of vision disturbance may be less obvious stroke symptoms for the patient. These symptoms may be recognized by non-health personnel in only severe cases, complicating recognition and clarification during the emergency call. As gaze deviation is a relevant symptom when assessing for large vessel occlusion possibly eligible for EVT, we believe that visual symptoms in the EMCC setting requires further study.^34^

Among the operational factors, we found that wake-up strokes were more likely to be suspected as strokes by the EMCC compared with non-wake-up strokes. We have not identified any research on wake-up strokes and EMCC, highlighting this as an area of interest for further research.

We performed a manual and time-consuming review of the 1,164 pathways of patients with stroke through the prehospital system. For future studies, we recommend the use of an automated system that uses both the dispatch protocol criterion code and text written by the medical dispatchers to calculate the accurate EMCC sensitivity. In cases of uncertainty, the audio log from the emergency call should be used to determine whether a stroke was suspected.

This study has some limitations. The dataset in this study contains no information on risk factors, such as prior stroke, hypertension, and diabetes mellitus. However, this aligns with the reality faced by most medical dispatchers, who often lack the time to access patient records in emergencies. To improve rapid stroke call decisions, systems that provide an easily accessible overview of a patient’s medical history are needed. Audio logs from the emergency calls were available for only the 2019 study population. Upon extraction, we found that the 2018 patient logs had routinely been deleted due to a data security policy. However, since most patients had a clearly defined prehospital route based on the structured data, only the audio logs were needed in a few cases.

Despite the limitations above, the quality of the NSR data is validated and considered a reliable source for healthcare studies.^35^ The data used encompassed an entire catchment area over a predefined period, leaving little reason to believe that patients with stroke were not identified. However, since the NSR data represent hospital records, the patient’s condition may have changed between the time of the medical emergency call and examination in the emergency room. All patient cases are manually reviewed, resulting in a unique dataset that provides substantial material suitable for addressing this study’s aims. Larger multi-center studies on the topics highlighted in this study could address the limitations presented in this study.

In conclusion, this study presents a novel and more accurate method for calculating EMCC stroke sensitivity. We propose adapting this method in future research, especially when investigating and comparing the accuracy of EMCC decision support tools, such as AI. The technique can be generalized to various diagnoses and EMCC systems in other regions and countries. Further, we have identified factors of particular interest for future EMCC research relevant to developing AI decision-support tools.

## Data Availability

All data is saved on a secure, restricted access research server at Haukeland University Hospital. Anonymized data can be made available upon reasonable request.

## Acknowledgments

The authors thank Bjørn Jamtli (Oslo) and Erlend Hodneland (Bergen) for their valuable input to the project and Merethe Landaas for representing the patients’ perspective. We thank Editage [http://www.editage.com] for editing and reviewing this manuscript for the English language.

## Author’s contribution

GB and EI researched the literature, conceived the study, and developed the protocol in collaboration with ASL and CA. EI wrote the first draft of the manuscript, obtained ethical approval, and was responsible for all patient contact. LM, CA, KKH, and EI extracted data and performed the statistics. All authors reviewed and edited the manuscript and approved the final version.

## Supplemental material

Variables from The Norwegian Stroke Registry

Variables from the emergency medical communication center

List of non-stroke-related chapters in the Index assigned to patients with stroke STROBE checklist

## Non-standard abbreviations and acronyms

AI: artificial intelligence
AMIS: Emergency Medical Information System
AISMEC: Artificial Intelligence Support in Medical Emergency Calls
EMCC: emergency medical communication centers
EPCC: emergency primary care center
EVT: endovascular thrombectomy
FAST: Face – Arm – Speech – Time
IVT: intravenous thrombolysis
LR: logistic regression
NIHSS: National Institutes of Health Stroke Scale
NSR: Norwegian Stroke Registry
OR: odds ratio

